# Swiss Physicians’ Experiences Of Sexual Harassment At Work. A Nationwide Survey

**DOI:** 10.64898/2026.10.19.26365908

**Authors:** Kevin T.J. Dzi, Patrick Bodenmann, Ralf J. Jox

**Affiliations:** Department of Education and Research (DFR), Institute of Humanities in Medicine, Lausanne University Hospital and University of Lausanne, Lausanne, Switzerland; Department of Vulnerabilities and Social Medicine, Unisanté, Lausanne, Switzerland

**Keywords:** sexual harassment, discrimination, physicians, survey, healthcare institutions

## Abstract

**Background:** Workplace discrimination and violence against health workers including sexual harassment is a societal problem worldwide. We aimed to assess the frequency, predictive factors, and effects of sexual harassment using a nationwide survey of physicians working in Switzerland.

**Materials and Methods:** We conducted a self-administered questionnaire survey of physicians working in Swiss healthcare institutions. The questionnaire assessed experiences of sexual harassment at work, including frequency, types, and sources of harassment. It also asked about the impact of sexual harassment, whether incidents were reported, and what actions were taken following a report.

**Results:** Among the 1837 physicians that participated in the survey, 575 (31.3%) had experienced sexual harassment at least once in their job. Among those reporting sexual harassment, 39.1% of women and 13.2% of men had experienced harassment at least once in their job. Women reported more verbal harassment, while men reported more physical harassment. Women were also more likely to face both forms combined. The main sources of harassment were colleagues of the same profession (46.7%), patients (42.3%), and superiors (41.0%). The most frequently reported impacts included stress (60.1%), emotional exhaustion (34.7%), reduced motivation at work (30%), and considering leaving the job (7.5%). Gender-men was associated with a lower likelihood of experiencing harassment 0.24 (0.18 to 0.31), whereas working in the German-speaking region was associated with a slightly higher likelihood of having experienced sexual harassment at least once (OR 1.35, 95% CI 1.02-1.81).

**Conclusions:** Sexual harassment is common among physicians in Swiss healthcare institutions. It adversely affects the mental health and career trajectories of both men and women yet remains widely underreported due to fears of reprisals and inaction. Institutions should effectively intervene to ensure that reported incidents result in appropriate actions, and that those who report them receive adequate protection from reprisals.

## Background

The wellbeing and performance of health care professionals are negatively impacted, among others, by institutional and workplace factors such as long working hours, involvement in medical errors, discrimination, and sexual harassment [1–4]. In this article, we focus on one key factor that can diminish healthcare professionals’ wellbeing and thus may indirectly also affect quality of patient care: sexual harassment. Sexual harassment includes unwanted intrusive physical acts ranging from unsolicited touching up until rape, as well as verbal behaviour like derogatory jokes, threats, obscene language, sexual propositions, or sharing offensive material [4]. Such actions create a negative psychological and emotional work environment that may affect the health of the person being targeted and lower quality of life [4, 5]. Sexual harassment experienced by healthcare workers is increasing globally, and although most studies focus on nurses and physicians [4, 6, 7], it affects health care workers from different professions such as midwifery [8] and medical laboratory [9], amongst others. Medical students may also encounter sexual harassment during their training [10]. In the healthcare setting, sexual harassment can be perpetrated by patients, their relatives, and other professionals, especially senior coworkers and superiors [11, 12].

A 2025 survey of physicians which involved 4339 doctors from 6 European countries revealed that over the course of 3 years, 6% of doctors in France, 5% in Germany, 5% in Italy, 3% in Portugal, 5% in Spain and 9% in the UK had personally experienced sexual abuse, harassment, or misconduct at work [13]. A 2019 study by Nagger and colleagues in the French-speaking region of Switzerland assessed whether 889 medical students and physicians had ever experienced gender discrimination and sexual harassment in the course of their work. Both men (16.7%) and women (19.0%) reported similarly high rates of sexual harassment [14]. Recently, a 2024 national survey on sexual harassment at the workplace conducted by the Swiss State Secretariat for Economic Affairs and the Swiss Federal Office for Gender and Equality revealed that among the 2553 employees from different sectors who completed the survey, one in three of the employees had experienced sexual harassment at work at least once, with a clear gender imbalance: 44% of women compared with 17% of men report such experiences. Jobs involving frequent customer contact such as hospitality, banking, and healthcare show particularly high exposure [15].

In Switzerland, campaigns against sexual harassment in both hospital settings and other sectors are regularly conducted to raise awareness among healthcare workers and the general public, and most health institutions have policies addressing sexual harassment. By reporting stories about sexual harassment and helping to raise awareness, the media also plays an important role [16, 17]. In addition, the Federal Office for Gender Equality promotes awareness campaigns and supports research on this issue. Article 328 of the Swiss Code of Obligations states that employers have the obligation to protect their employees from sexual harassment and also ensure that their employees don’t suffer further adverse consequences [18]. On a more general level, article 4 of the Swiss Gender Equality Act explicitly prohibits sexual harassment as a form of discrimination, including behaviour such as threats, coercion, pressure, or offering advantages in exchange for sexual favours [19].

As seen in the various studies above, although physicians are often perceived as occupying a higher position within the healthcare hierarchy, they are not exempt from experiencing sexual harassment. Such harassment carries significant professional and personal consequences, paralleling the effects of other forms of discrimination. These impacts may include reduced enthusiasm for work, difficulties with concentration, emotional exhaustion, burn out and, in some cases, decisions to leave their careers altogether [20, 21].

National studies examining sexual harassment among physicians from all specialities are sparse, in Switzerland as in other countries. The present study sought to address this gap by assessing the prevalence, types, sources, reporting behaviours, and consequences of sexual harassment within the medical profession in Switzerland. In addition, the study examined the predictive factors associated with these outcomes to provide a more comprehensive understanding of the phenomenon.

## Methodology

### Study design and study population

We conducted an anonymous, cross-sectional, self-administered online survey among physicians working in Switzerland from June to August 2025. The survey was open to physicians of all specialities, both resident physicians in specialty training and those with board certificates. Ethical approval was obtained from the Institutional Review Board of the Faculty of Biology and Medicine of the University of Lausanne (project No: E-FBM-042024-00001). Prior to participating in the survey each participant was required to sign an online consent form.

### Data collection

The semi-quantitative questionnaire was part of a larger survey on work-related discrimination experienced by physicians and nurses, designed by the authors KD and RJJ. The instrument was created based on previous studies assessing the discrimination experiences faced by physicians and nurses and as well as studies that measured stress and burnout related to work-related discrimination [22–28]. The items used in these studies had been validated; however, authors sometimes reformulated questions to suit their specific research objectives. For example, in the study by Padela and colleagues[27], the question on discrimination used by Nunez and colleagues [28] was modified from assessing the experience and frequency of workplace discrimination over one’s career to assessing the experience and frequency of religious discrimination over one’s career. We did the same while developing our questionaire, using certain items unaltered as presented in their original studies while adapting others to suit our reseach objectives. For example, the questionnaire used in the Swiss German part to measure discrimination included 10 forms of discrimination or bullying that participants could select from [24]. In our questionnaire, we retained all of these items but excluded those relating to bullying, mobbing, and violence, given our focus on discrimination. The same items employed to assess burnout and stress in their survey were utilised in our study as well. To measure the frequency of experienced discrimination, we used the same question as Dyrbye and colleagues[22]; however, the response categories were slightly modified. In their study, participants could choose from the categories “never,” “once,” “several times a year,” “weekly,” and “several times a week.” In our study, these categories were adapted to “once,” “several times a day,” “several times a week,” “several times a month,” and “several times a year.”

The part on sexual harassment was developed in collaboration with the Swiss Association of Resident and Attending Physicians (VSAO-ASMAC) and was tailored specifically to physicians. While some questions were adapted from existing surveys cited above [27, 28] others were developed denovo. The latter were primarily open-ended items, such as those asking participants to describe the type of harassment experienced, actions taken following reporting, and to whom the harassment was reported. The questions adapted from the surveys mentioned above included “Have you ever experienced religious discrimination in your career?”, which was modified to “Have you ever experienced sexual harassment at least once in your work?”. In addition, the question on the “frequency of experiencing religious discrimination” was adapted to “frequency of experiencing sexual harassment”. The question on consequences was modified from “Have you ever left a job (as a physician) due to encountering discrimination in your workplace?” to a multiple-response question allowing respondents to select one or more consequences they had experienced as a result of sexual harassment. To ensure that the questionnaire was understandable and appropriate, we pilot-tested it with six physicians and two nurses working at different Swiss hospitals. Their feedback was used to modify the questionnaire and develop the final version.

The whole questionnaire was composed of 6 parts: I) sociodemographic data, II) experiences of all forms of discrimination, including the sources, perpetrators, consequences, reasons for reporting/not reporting, III) experiences of sexual harassment including types, sources, description of harassment, consequences, frequency and action taken when harassment occurs IV) attitudes on discrimination, elicited by using vignettes V) measurement of personal and workplace burnout. For the purpose of this article, we focus on the questions from sections I & III. From section I, sociodemographic data collected included gender, age group, migratory background, continent of origin, citizenship, continent of origin of mother and father, years of professional practice, the speciality, and the work context. In section III, the main question was whether the participant had experienced sexual harassment at least once in their job. Additional questions for those who had experienced sexual harassment included types of harassment experienced (physical, verbal, both), description of the type of harassment experienced, the frequency of harassment experienced (here only frequency in the past 12 months was considered), the sources of harassment, the consequences of experiencing harassment, action taken when harassed (reporting/person harassment incident was reported to), and reasons for non-reporting. The questionnaire was developed in English and translated into the three main languages of Switzerland, German, French, and Italian, by native speakers of the respective languages. To conduct the online survey, we used the open-access platform Qualtrics, and data was collected anonymously. The VSAO -ASMAC distributed the survey to its members via its monthly newsletter, which is sent to all 24’318 registered members, and also shared it on its LinkedIn account. Reminder messages were subsequently sent via email and LinkedIn by the VSAO-ASMAC. Survey data was handled in a confidential manner, and only authors KD and RJJ had access to data that was stored on a secured server at the Lausanne University Hospital.

### Statistical analysis

Sociodemographic characteristics were summarised according to whether respondents reported having experienced sexual harassment at least once in their job, as opposed to those who had not experienced it or declined to answer. Descriptive plots were generated for selected categorical variables with a large number of categories, such as physician speciality. Continuous variables, including years of professional experience, were summarized using means, standard deviations, medians, and interquartile range. Statistical associations between sociodemographic variables and the outcome variable *“Have you ever experienced sexual harassment at least once in your job?”* were assessed using the chi-square test or Fisher’s exact test where appropriate. Multivariable logistic regression was used to examine the association between sociodemographic characteristics and having experienced sexual harassment at least once on their job (outcome variable), while adjusting for relevant covariates, including gender, age group, migration background, region of practice, and work context. For this analysis, participants who selected “no answer” to the question on lifetime experience of sexual harassment (n = 40) were excluded. We further excluded participants who reported their gender as “other” or “prefer not to answer,” as well as those with missing information on migration background (n = 23). Additionally, respondents from the Suisse Romansch region (n = 5) and those aged 18-24 years (n = 4) were excluded due to small numbers and the absence of reported harassment in these groups. The age group ≥65 years was combined with the 55-64 years category due to small cell counts (3 reporting harassment and 24 not reporting harassment). The final sample included 1,765 participants for the logistic regression analysis. A p-value of less than 0.05 was considered statistically significant. All analyses were conducted in R version 4.3.0. Reporting of the survey follows the CROSS-reporting guideline [29]. It should be noted that the sample size for each analysis is reported separately in the results section. Denominators vary across analyses due to item-level missing data and because certain analyses were conducted in relevant subgroups only.

## Results

### Study sample

There were 1837 physicians who participated in the survey. Their sociodemographic characteristics are summarized in table 1 in the first column. More than two third of the respondents were women (70.2%) and more than two thirds had an age between 25 and 44 years (67.9%). While a bit more than half of the respondents had Swiss citizenship (57.2%), about half of the responding physicians indicated having a migratory background (50.7%). More than 90% indicated Europe as their continent of origin. About half of the sample identified themselves with the Christian religion and 40% with no religion. Three quarters of the respondents work in the German-speaking part of Switzerland, 17.5% in the French-speaking part, and 2.7% in the Italian-speaking part. Three quarters of our sample pursue their professional activity in a hospital, and their mean professional experience as physicians is about 12 years.

**Table 1.**
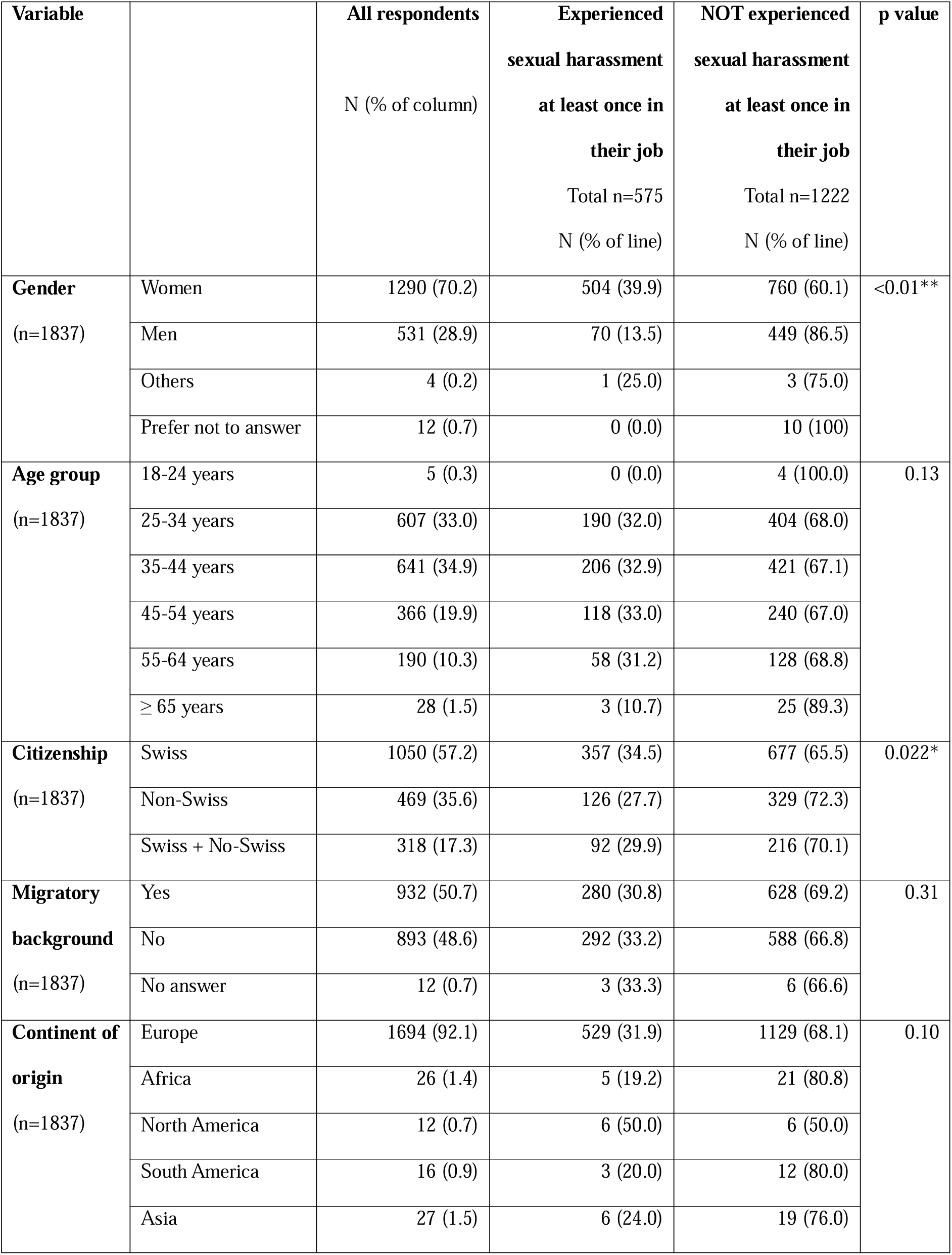
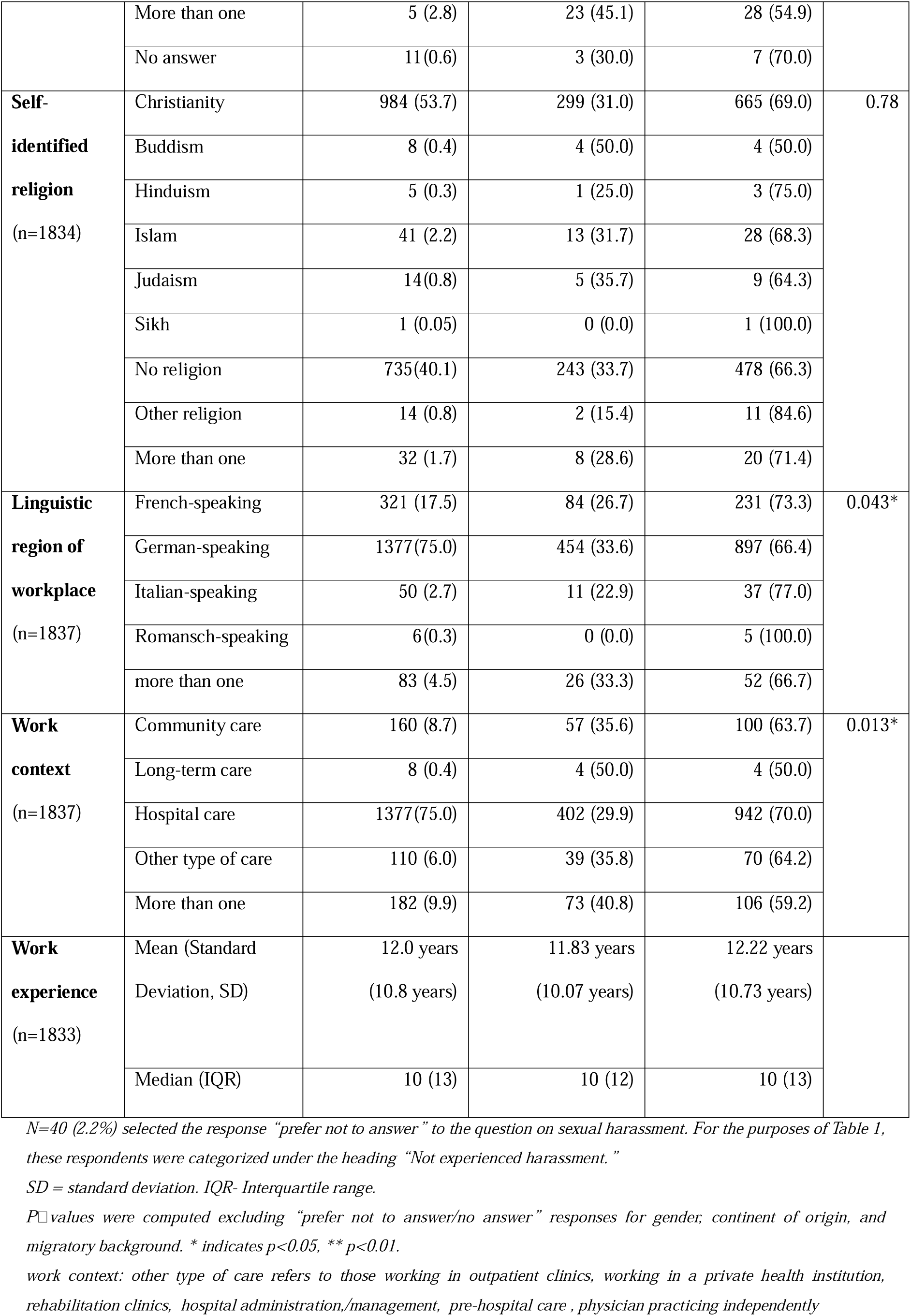
Sociodemographic characteristics and frequency of reported sexual harassment (n=1837)

### Prevalence of sexual harassment

Among the respondents, almost one third (31.3%) reported having experienced sexual harassment at the workplace at least once in their job. The proportion of women who reported experiencing sexual harassment at least once in their job was three times higher than that of men, a finding that was statistically significant (Table 1). Across the age groups of 25-34, 35-44, 45-54, and 55-64 years, the prevalence of reported sexual harassment, experienced at least once in their job, was relatively consistent at approximately 31-33%. In terms of work context, higher rates of were reported by those who practiced in more than one context and those who practiced in long term care settings compared to the other settings. When considering the linguistic region of their workplace, individuals who practiced in more than one region and those working in German-speaking Switzerland reported slightly higher rates of having experienced sexual harassment at least once in their job. Moreover, participants who identified as Swiss reported experiencing sexual harassment at least once in their job more often than those who identified as non-Swiss or as both Swiss and non-Swiss.

Regarding medical specialties, physicians working in surgical fields, internal medicine, family/general medicine, orthopedics, and obstetrics and gynecology reported higher rates of sexual harassment compared to those in other specialties (Fig 1). The large specialties where we had samples of n>100 all show proportions of more than 30% of respondents having experienced sexual harassment, with surgery attaining 38.0%. Only specialties with at least 10 respondents are shown in the figure.

**Fig 1.**
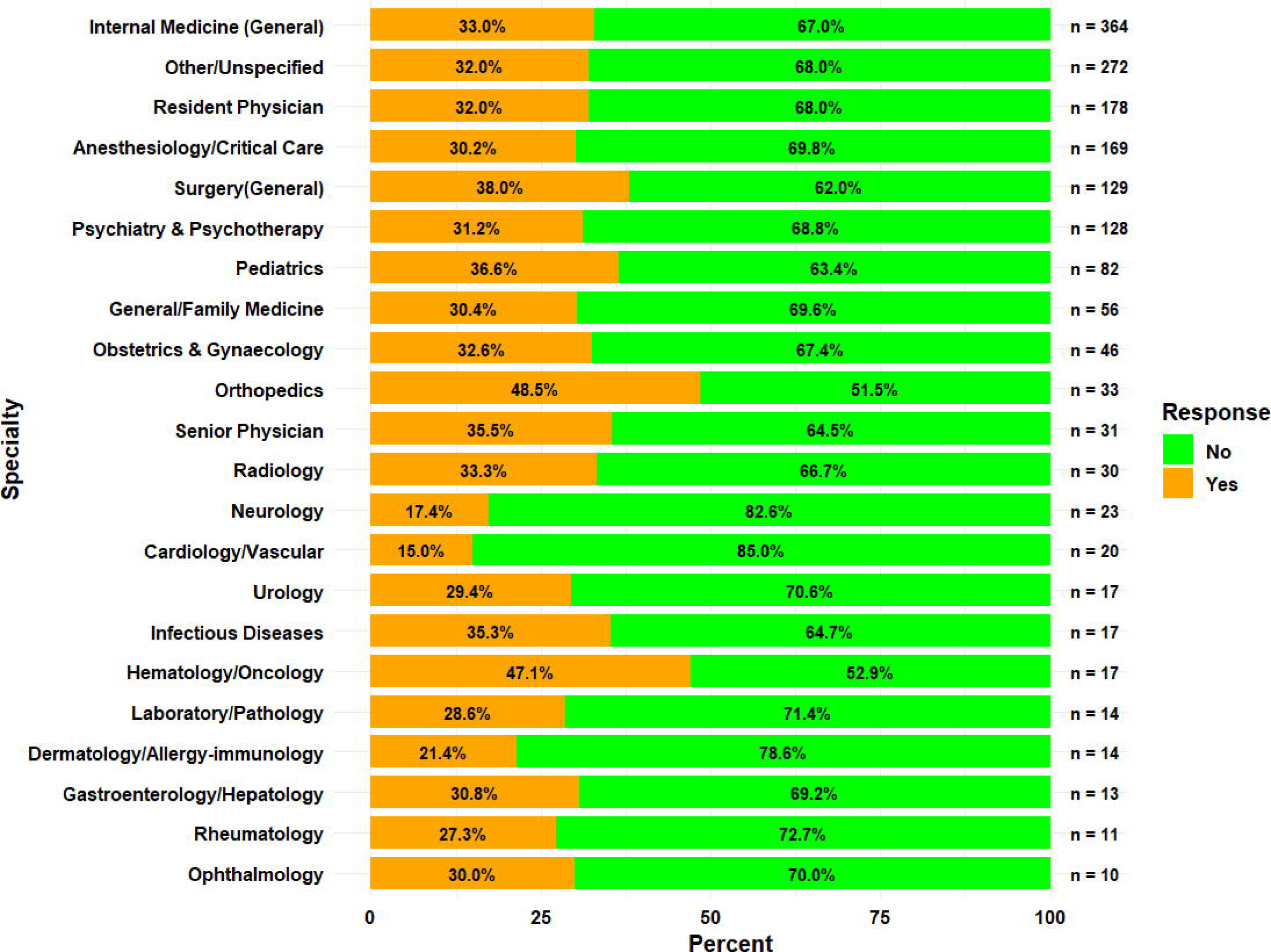
Specialities and experience of sexual harassment (n=1674) The figure shows the percentage of respondents who reported having experienced sexual harassment at least once in their job, differentiated according to the medical specialty they work in.

### Frequency of sexual harassment

Those who indicated having experienced sexual harassment at least once in their job were also asked how often they experienced it in the past 12 months. While 313 (57.5%) reported that they had experienced sexual harassment once during this period, 184 (33.8%) reported a frequency of several times a year, 31 (5.7%) several times a month, 10 (1.8%) several times a week, and 6 (1.1) several times a day. When examining whether the frequency of experiencing sexual harassment varied across sociodemographic variables, younger participants (25-54 years) reported harassment across all frequency categories, whereas older participants (55+ years) were concentrated in the ‘once’ or ‘several times a year’ categories (Table 2). The two respondents from the oldest age group (≥65 years) reported experiencing harassment on a daily basis.

**Table 2.**
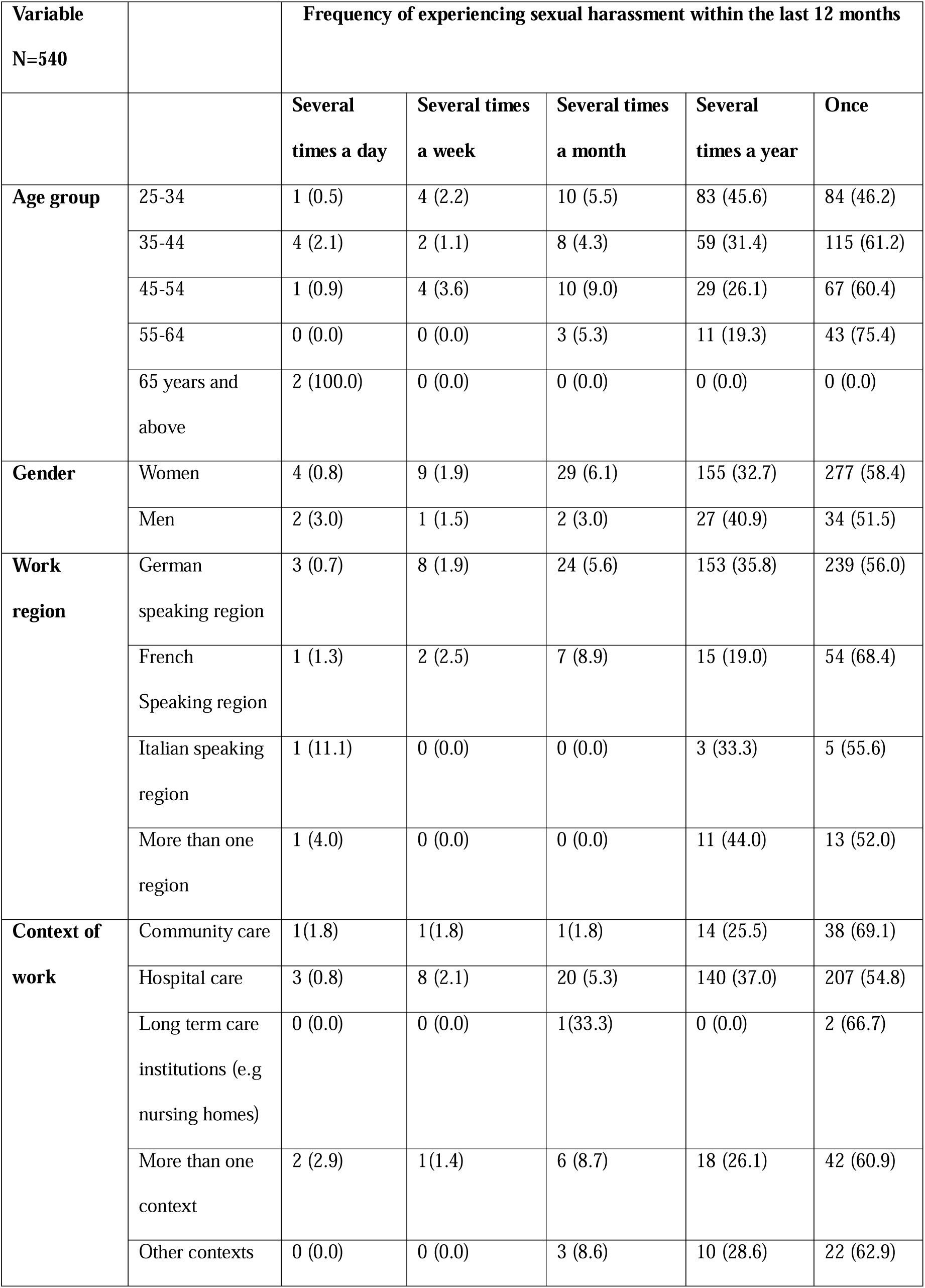
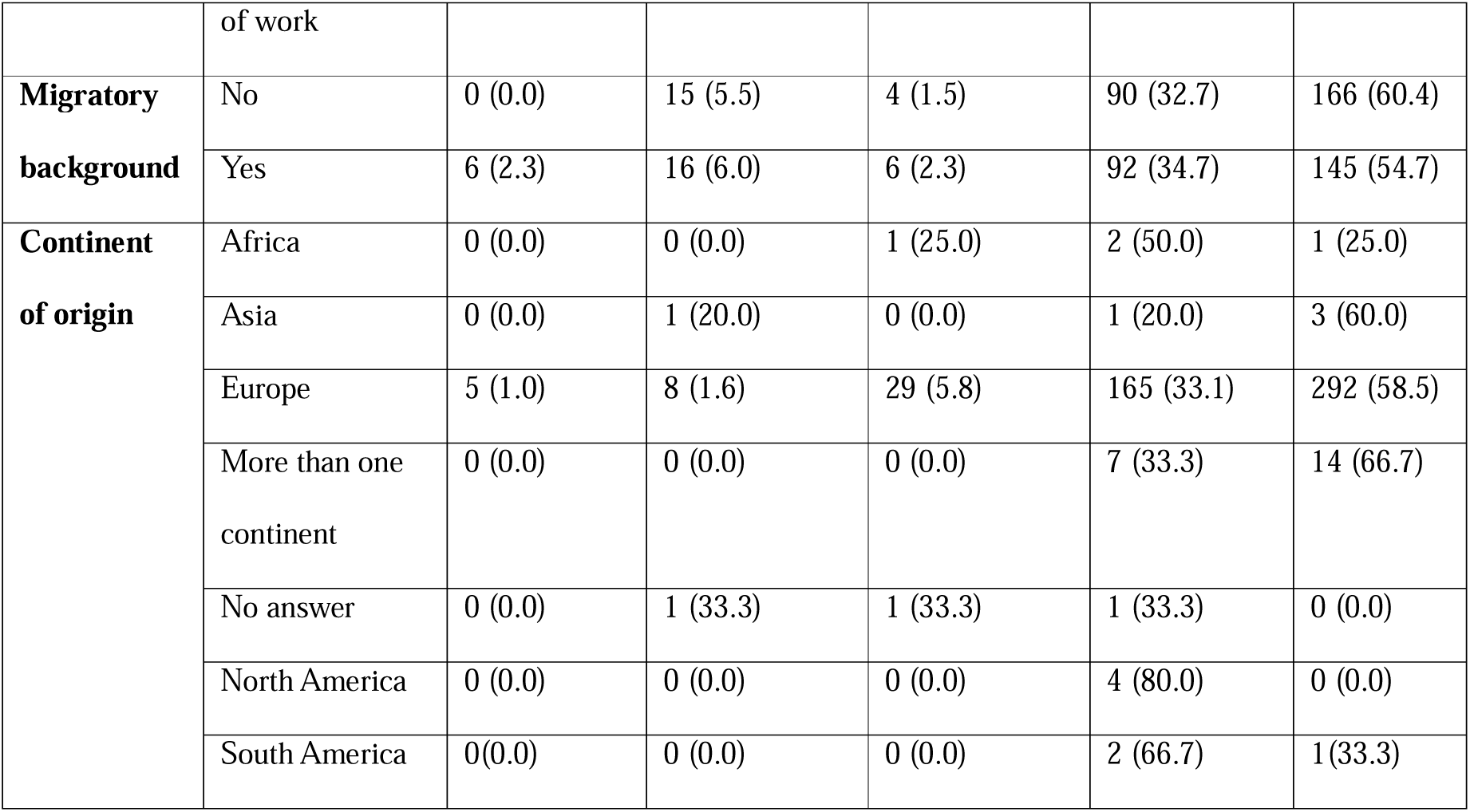
Distribution of sexual harassment frequency by socio-demographic characteristics.

Across all linguistic regions, the majority of the participants reported experiencing sexual harassment once or several times a year, with relatively few reporting experiencing harassment several times a week or several times daily. The distribution of frequencies appeared similar across the German-, French-, and Italian-speaking regions, indicating no clear regional differences at the descriptive level. For gender, work context, and migratory background, the distribution of frequencies also appeared similar across the different subcategories of these variables, with most participants reporting having experienced sexual harassment only once and very few reporting experiencing harassment several times weekly or several times daily.

### Multivariable logistic regression analysis

In the multivariable logistic regression analysis, men gender was significantly associated with having a lower likelihood of having experienced sexual harassment at least once in their job (OR: 0.24, 95% confidence interval, CI, 0.18-0.31), (p<0.01) (Table 3). Participants who work in the German-speaking part of Switzerland had a higher likelihood of having experienced sexual harassment at least once in their job compared to participants in the French-speaking part (OR: 1.35, 95% CI: 1.02 to 1.81), (p=0.039). Having a migratory background was not a significant independent predictor of this outcome. Those working in long-term care facilities were more likely to have experienced sexual harassment at least once in their job compared to those working in community care, but the association failed to reach statistical significance after adjustment. The logistic regression model was statistically significant, ^2^ (12) = 146.47, p < .001, and demonstrated modest discriminatory capacity (AUC = 0.66), indicating a limited ability to distinguish between individuals who did and did not experience sexual harassment at least once in their job. The Hosmer-Lemeshow test suggested no evidence of lack of fit (p = 0.73), indicating that predicted probabilities were broadly consistent with observed outcomes. However, the model’s explanatory power was relatively low (Nagelkerke R^2^ = 0.11), suggesting that important structural, cultural, and contextual factors shaping experiences of sexual harassment may not be fully captured by the variables included.

**Table 3.** Association between sociodemographic characteristics of physicians and the reported experience of sexual harassment at least once in their job (n=1765)

| Sociodemographic factor | OR (95% CI) | P - value |
| --- | --- | --- |
| Gender-women | - | - |
| Gender-men | 0.24 (0.18 to 0.31) | P<0.01* |
| Agegroup25-34 | - | - |
| Agegroup35-44 | 1.1 (0.85 to 1.42) | 0.46 |
| Agegroup45-54 | 1.05 (0.78 to 1.41) | 0.75 |
| Agegroup≥55 | 0.91 (0.62 to 1.32) | 0.63 |
| Migratory background: No | - | - |
| Migratory background: Yes | 0.97 (0.79 to 1.20) | 0.77 |
| Context: Community care | - | - |
| Context: More than one context of work | 1.36 (0.86 to 2.17) | 0.19 |
| Context: Hospital care | 0.83 (0.58 to 1.19) | 0.30 |
| Context: long term care facilities | 1.92 (0.42 to 8.89) | 0.39 |
| Context: other contexts | 1.13 (0.66 to 1.93) | 0.66 |
| Region: French-speaking region | - | - |
| Region: German-speaking region | 1.35 (1.02 to 1.81) | 0.039* |
| Region: Italian-speaking region | 0.87 (0.40 to 1.79) | 0.71 |
| Region: More than one region | 1.45 (0.82 to 2.52) | 0.20 |

\*significant

### Types and occurrence of sexual harassment (n=544)

We also strived to differentiate the various common types of sexual harassment which physicians had experienced in their job. Among the 544 respondents who answered this question, 300 (55.1%) had experienced verbal sexual harassment, 73 (13.4%) physical sexual harassment, and 171 (31.4%) both forms, physical and verbal sexual harassment. Gender-based differences were observed in the types of harassment reported by participants. Among men, the proportion of physical sexual harassment reported was higher (24.2%, n=16 out of 66) than among women (11.9%, n=57 out of n=477). In contrast, verbal harassment was slightly more prevalent among women (55.3%, n = 264 out of 477) than men physicians (53%, n = 35 out of 66). Women also reported a higher occurrence of combined physical and verbal harassment (32.7%, n = 156) relative to men (22.7%, n = 15). A chiLsquare test demonstrated that the association between gender and the type of harassment experienced was statistically significant (χ² = 8.41, df=2, p=0.015).

Qualitative free-text responses provided more detail on the types of sexual harassment experienced. Some examples narrated by women physicians include: “*Drunk/intoxicated patients in the emergency room regularly harass me verbally and physically in a sexual manner. Verbally with suggestive comments and physically with touching*”; “*Advances from superiors; threats of having to perform rectal examinations if I didn’t give the right answer; daily comments about my appearance; humiliation during meetings: as a woman, you’ll never be a doctor in our department…in short, the list is too long!”*; “*Experienced occasional unwanted advances from male patients*”; “*Comments on my sexual experience, my physical appearance, superiors telling their sex stories to me in detail*”. Examples of some types of sexual harassment highlighted by men physicians include: “*I was touched inappropriately several times by female assistant doctors standing on the same side of the operating table*”; “*Inappropriate comments from female patients*”; “*Touching of the buttocks by female patients*”; “*I was inappropriately touched by a male nursing staff member in the locker room*”.

### Sources of sexual harassment

Physicians experience harassment from multiple sources in healthcare institutions. Table 4 presents the agents of sexual harassment according to the responding physicians. Colleagues of the same profession, patients and superiors were the most often reported perpetrators. 46.7% of physicians reported that the sexual harassment they experienced originated from colleagues within the same profession, while 42.3% identified patients as the source. Additionally, 41% attributed it to superiors, and 20% to colleagues from different professions. A higher proportion of men physicians than women physicians reported experiencing harassment from administrative staff and colleagues from other professional groups.

**Table 4.** Perpetrators of Sexual Harassment. (multiple responses allowed) (n = 544)

| Agents of sexual harassment | Reported frequency (all victims of harassment)<br>N (%) | Reported frequency: Women victims N(%) | Reported frequency: Men victims<br>N(%) |
| --- | --- | --- | --- |
| Colleagues of the same profession | 254 (46.7) | 233 (48.8) | 21 (31.8) |
| Patients | 230 (42.3) | 204 (42.8) | 25 (37.9) |
| Superiors | 223 (41.0) | 209 (43.8) | 14(21.2) |
| Colleagues of different professions | 109 (20.0) | 87 (18.2) | 22 (33.3) |
| Relatives or friends of patients | 42 (7.7) | 38 (8.0) | 4 (6.1) |
| Administrative personnel | 20 (3.8) | 14 (2.9) | 6 (9.1) |
| Other (please specify)<br>* | 14 (2.6) | 10 (2.1) | 4 (6.1) |

\*Others: Anaesthesia nurse; midwife, teacher, nursing management, patient surrogate, supervisor, bystanders

### Consequences of sexual harassment

Among the 544 respondents who reported consequences of sexual harassment, the most commonly self-reported consequence was stress, affecting 60.1% of respondents. Other significant sequelae included emotional exhaustion (34.7%) and lack of motivation (30.0%). Additional effects reported were lack of concentration (22.2%), absenteeism (5%), and leaving the job (7.5%). Among the 477 women who experienced harassment, 62.7% reported being stressed while 42.4% out of the 66 men who experienced harassment reported being stressed. Also, a slightly higher proportion of women who experienced harassment reported lacking motivation at their job compared to men (see Fig 2).

**Fig 2.**
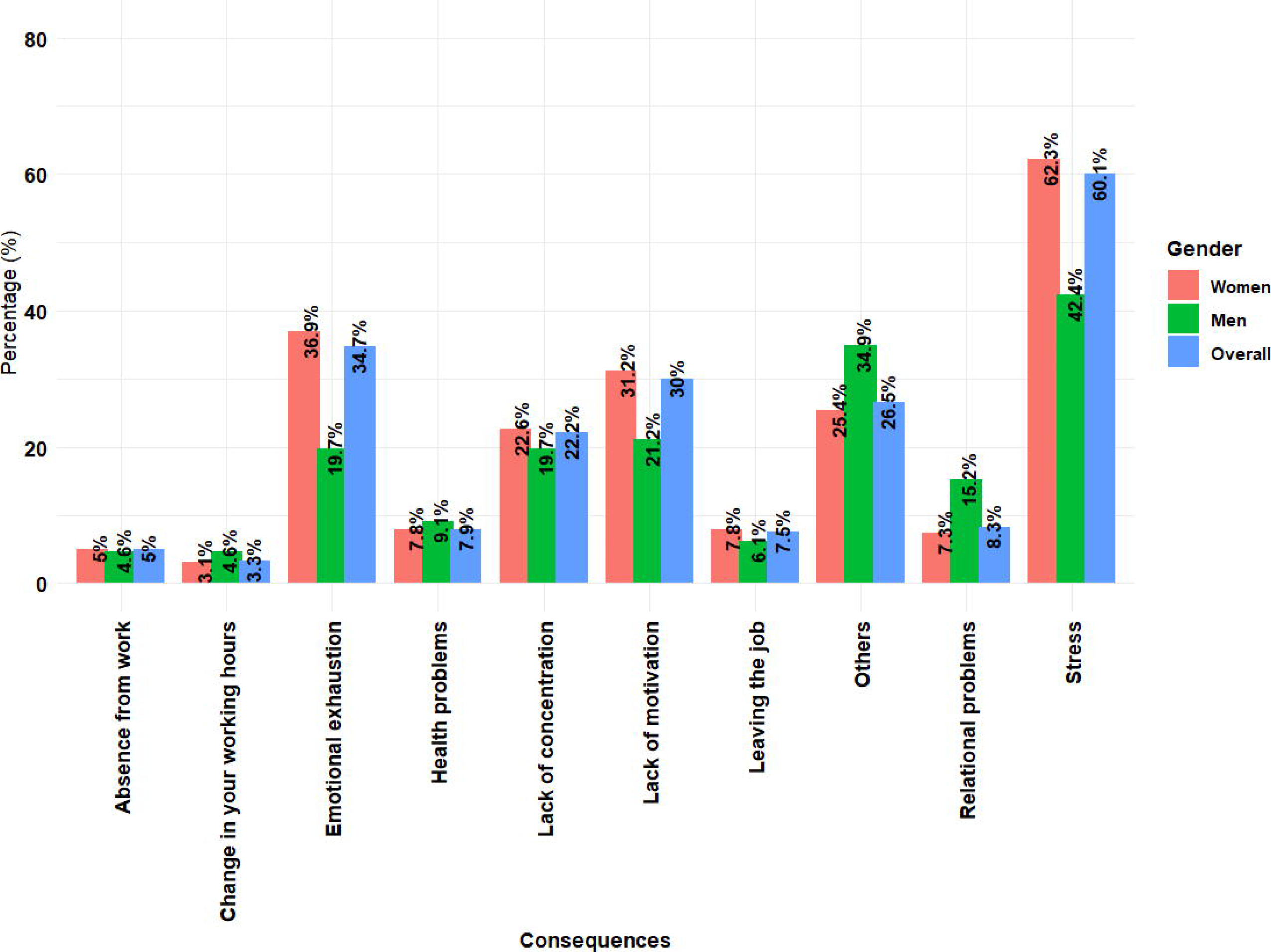
Consequences of experiencing sexual harassment (multiple responses allowed) (n=544) Other responses in the free-text field: feelings of anger, frustration, and powerlessness, losing trust in superiors (particularly when they were the perpetrators), fear of working with colleagues involved in harassment, sleep disturbances, disappointment, sense of injustice.* The overall number of respondents (n=544) included women=477, men= 66 and Gender-other =1

### Reporting sexual harassment

Those who experience sexual harassment may or may not report their experience. Among the 544 respondents who answered this question, 386 (71%) indicated that they did not report sexual harassment when they experienced it, whereas 158 (29%) did. When differentiating according to gender, 30.2% of the women (143/474) who experienced sexual harassment chose to report it, compared with only 21.2% (14/66) of men who reported their experience. Reporting patterns also varied across age groups: younger cohorts were more likely to come forward than older ones. Among individuals aged 25-34, 32.4% (59/182) reported harassment, followed by 31.4% (59/188) in the age group between 35-44 years, 27% (30/111) in the 45-54 years group, and just 14% (8/57) among those aged 55-64 years.

When asked to whom they reported incidents of sexual harassment, the descriptive responses provided by 152 participants indicated that most physicians reported their experiences to supervisors, department heads, or the chiefs of service. Others directed their complaints to coworkers, human resources, hospital administration, or organizations addressing sexual harassment. Furthermore, when participants were asked whether the individuals they reported to took action, 97 of 155 respondents (62.6%) confirmed that action was taken, whereas 58 (37.4%) stated that no action was taken. Free-text answers revealed the following types of actions taken: addressing the perpetrator directly, facilitating mediation between the perpetrator and the victim, offering support by supervisors, documenting the incident, encouraging the victim to make the case public, but also advising the victim to tolerate it, especially when the perpetrator was a senior colleague. When sexual harassment was authored by a patient, the types of actions mentioned were sending another professional to the patient to prevent further harassment, refusing to care for the patient or discharging the patient (see S1 Table for detailed free text responses).

### Non-reporting of sexual harassment

The participants who chose not to report the sexual harassment they experienced provided a range of reasons for this decision (see TableL5). Overall, the most common reason cited by nearly twoLthirds of respondents was the belief that reporting would not lead to any meaningful action. A slightly higher proportion of men than women indicated that they did not know whom to report to. In contrast, nearly twice as many women as men reported that they avoided reporting the harassment because they did not want to be perceived as weak, attention-seeking, or as victims (Table 5).

**Table 5.** Reasons for not reporting sexual harassment (multiple responses allowed) (n=381)

| Reasons for not reporting | Frequency<br>Overall<br>(n=381)<br>N (%) | Frequency Women<br>(n=330)<br>N(%) | Frequency<br>Men (n=51)<br>N(%) |
| --- | --- | --- | --- |
| I did not think that reporting would result in any meaningful action | 243 (63.8) | 219 (66.4) | 24 (47.1) |
| I did not wish to be perceived as weak, attention seeking, or as a victim | 115 (30.2) | 106 (32.1) | 9 (17.6) |
| I felt embarrassed, fearful or ashamed | 106 (27.8) | 93 (28.2) | 13 (25.5) |
| I did not know who to report to | 104 (27.3) | 88 (26.7) | 16 (31.4) |
| I did not feel supported by my peers, supervisors or the institution | 101 (26.5) | 89 (30.0) | 12 (23.5) |
| Other reasons written in free text field | 112 (29.4) | 90 (27.3) | 22 (43.1) |

Additional reasons for not reporting were mentioned in a free-text field indicated in Table 5 above: fear of retaliation (especially when perpetrators are employers or superiors); previous offenses had occurred in the past without consequences, minimizing harassment if it was only verbal or considered too minor to report; they had acquired some training on managing sexual harassment situations so managed it themselves; they decided to confront perpetrators of sexual harassment themselves; fear that reporting will harm their reputation, fear that they may not be believed (especially if victim was a man), not being sure if the incident constituted sexual harassment, lack of formal reporting system, no witness available.

## Discussion

This study presents the first nationwide survey on sexual harassment encountered by physicians across several specialities in Switzerland. Overall, 31.3% of the responding physicians reported having experienced sexual harassment at least once in their job. The frequency was 39.1% among women and 13.1% among men, indicating a significant gender difference in terms of becoming victim of this kind of behaviour. The main sources of harassment reported in our study included patients, colleagues of the same profession and superiors, and self-reported outcomes associated with experiences of harassment included diminished motivation, stress, emotional exhaustion, and other complications such as anger, frustration, and sleep disturbances.

The frequency of sexual harassment shown for men and women in our study are slightly lower than the published prevalence of workplace-related sexual harassment in the general population of Switzerland: according to a national study in 2024, 44% of women and 17% of men reported sexual harassment at work [15]. At the same time, the prevalence of sexual harassment reported by women physicians in our study is higher than that reported by Nagger and her colleagues in a 2019 study they carried out with physicians in the French-speaking part of Switzerland where women and men sexual harassment prevalence rates were 19% and 16.7% respectively [14]. Our finding that sexual harassment is much more prevalent among women physicians align with an Italian study examining sexual harassment and various forms of discrimination faced by healthcare workers, as well as multiple studies conducted on sexual harassment across different continents [20, 30].

Our findings show that sexual harassment is experienced in all major medical specialties from which we had high numbers of respondents, with a peak among those working in surgery. Similar patterns have been observed internationally. An observational study in the United Kingdom involving 756 surgical staff found that a majority of women (63.3%) reported experiencing sexual harassment by colleagues, along with a significant proportion of men (23.7%) [31]. It remains to be studied whether the particularly high prevalence in surgery observed in our study reflects selection bias, characteristics of the surgical work environment and culture, or a combination of these factors.

In our study, women reportedly experienced verbal harassment relatively more often than men, while men tended to report relatively more experiences of physical harassment than women. These findings are consistent with a study carried out among Swiss physicians working in internal medicine where verbal sexual harassment was reported by 74% women versus 38% men and physical harassment reported by 34% men versus 15% by women [32]. Yet, our findings reveal that, women had a higher percentage when asked if they had experienced both forms of harassment. This finding is in line with the results of the multivariable logistic regression analyses, which indicated that women had higher odds of experiencing sexual harassment after adjusting for other factors.

Furthermore, descriptive analyses revealed that one-third of participants from all age cohorts reported experiencing sexual harassment in the course of their work except for the eldest and youngest age categories (both had few respondents). This finding shows that sexual harassment is experienced by both younger and older physicians and measures need to be taken by institutions to protect their employees.

Cultural factors may also play a certain role. In multivariable regression analyses adjusted for age, gender, migration background, and work context, having the GermanLspeaking region as region of work was associated with a slightly higher likelihood of reporting sexual harassment at least once in their job compared with the FrenchLspeaking. However, this finding should be interpreted with caution, as other factors such as hierarchical position and organisational culture, which were not captured in our study, may influence sexual harassment experiences and reporting. Additionally, socio-economic background may affect reporting practices, and factors such as how previous harassment cases have been handled could influence reporting. In the 2024 Swiss survey among the general population mentioned above, no noticeable differences in sexual harassment rates between the language regions of Switzerland was found. A possible explanation is that their survey included individuals working across multiple sectors rather than solely within the health sector, whereas our study focused exclusively on physicians. In addition, their sample encompassed healthcare workers from a variety of professions, not only physicians, which may partly account for the differences in findings.

In terms of sources, the highest perpetrators of sexual harassment in our study were reported to be superiors, colleagues of the same profession, and patients. This is consistent with other national and international studies where perpetrators of harassment have both been colleagues and patients [4, 32]. The results are relevant since adequate prevention measures against sexual harassment in healthcare should be tailored to the different groups of perpetrators. While sexual harassment among physicians is amenable to intra-institutional measures (like sensitization, education, labour law responses), sexual harassment committed by patients entails other ethical implications and may require better communication, patient education, and public measures. In our study we also found that physicians employed strategies such as discharging the patient if they were the perpetrators. This has been recommended in other studies as a strategy for managing sexual harassment cases [33]. When sexual harassment originates from patients, it is of course ethically crucial whether the patient acted voluntarily or due to a mental condition, such as fronto-temporal spectrum disorders, addiction, psychosis, or delirium [34]. A strategy that has been suggested in some studies is that physicians preserve the professional physician-patient relationship by paying attention to their own verbal and nonverbal communication [33]. According to another study, a way of managing verbal harassment from patients is to redirect the conversation, ignoring the comment made by the patient, or to see patient as a team [34].

Research has shown that healthcare workers who have been victims of sexual harassment and other forms of discrimination suffer from reduced motivation, difficulty in concentrating, and changes in working hours that diminish the quality of life of healthcare workers and disrupt a healthy workplace environment[35, 36]. These findings are consistent with those observed in our study where one third of participants who experienced sexual harassment reported that they suffered from lack of motivation at work. Our findings align with those of the 2024 national Swiss survey across multiple employment sectors, where the most commonly reported negative consequences included feelings of shame, a deterioration of the work environment, and intentions to resign. Sexual harassment is a significant source of stress, as shown in our study, and as documented in other research can also contribute to burnout [37]. As reported in several U.S.Lbased studies, participants in our study also indicated that experiencing sexual harassment negatively affects their motivation at work and leads to stress, emotional exhaustion, and, in some cases, job changes [20]. It should be noted that sexual harassment could also break trust in the patient physician relationship leading to poor quality of care offered to patient [38].

Most physicians in our study who experienced sexual harassment did not report these events, which may be related to the finding that the perpetrators are often superiors. In the 2024 Swiss survey across multiple employment sectors only 18% were comfortable reporting to their supervisors; 51% preferred to report to their work colleagues while 57% preferred to talk about incident to persons in their private sphere[39]. Upon reporting sexual harassment to their superiors, our findings show that, while action was taken in some situations, in other situations, no action was taken. Descriptive texts from respondents provide examples of the types of actions taken, and it is evident that it was easier to act when the perpetrator was a patient than when it was a senior colleague or superior. This finding is consistent with a study from India, which showed that in hospital settings, patients who perpetrated sexual harassment were more likely to be held accountable/punished by superiors than physicians who committed similar acts [4].

The fact that a large majority of our participants choose not to report harassment is also consistent with international findings according to which the harasser is often in higher position of power and so the victim usually depends on the harasser for recommendations and promotions [40, 41]. Some physicians in our study indicated that they did not report because they felt it was not clear whether what happened actually constituted harassment. Measuring and monitoring sexual harassment is challenging because it is widespread and often underreported by those who experience it [42, 43]. Other participants in our study said they did not report because the confronted the perpetrator directly. This has also been reported in other studies as a coping mechanism [44, 45]. A major problem we identified is that some physicians did not report sexual harassment because they had observed in the past that reporting did not have any meaningful consequence, as also documented in the literature [40]. This is particularly problematic since it may lead to a vicious circle and reinforce patterns of sexual harassment behaviour as well as the underlying structural and cultural conditions.

## Limitations

Our study has limitations. One limitation is that the questionnaire focused only on two forms of harassment, verbal and physical, making it impossible to capture the prevalence of more subtle manifestations such as microaggressions. Nevertheless, the descriptive free-text responses provided insights into the broader range of sexual harassment experiences. A second limitation concerns the geographic distribution of participants. Most respondents were from the German-speaking regions, followed by the French-speaking regions, with very few participants from the Italian-speaking region, resulting in underrepresentation. This imbalance may be partly due to VSAO communications being primarily in German and French. We attempted to mitigate this by using snowball sampling to reach additional participants, particularly in Italian-speaking Switzerland. Finally, we were unable to calculate a response rate because the survey was disseminated through newsletters, social media channels such as LinkedIn, and snowball sampling. We chose this broad promotional strategy to reach as many physicians as possible and reduce the risk of underrepresentation. Another limitation of our study is the potential for sampling bias. Since the majority of participants were women physicians (70.2%) and most were aged 25-34 years (67.9%), the findings should be interpreted with caution when considering generalizability. The higher participation of women is not unexpected, as previous studies suggests that women healthcare workers are more likely to experience harassment in healthcare settings, which may also make them more willing to participate in studies on this topic. Also, a slightly higher proportion of the VSAO members are women. Nevertheless, some results were reported separately for both men and women participants to provide a balanced overview. Similarly, the greater representation of younger physicians may reflect the fact that early-career doctors are more frequently exposed to harassment compared to their senior counterparts as the distribution of VSAO members across different age groups remains fairly constant.

## Conclusion And Recommendations

### Conclusion

In conclusion, our study found that rates of sexual harassment experienced by Swiss physicians are high, affecting both men and women, although women are disproportionately impacted. These experiences have negative consequences for physicians, as victims in our sample reported suffering from increased stress, reduced motivation, emotional exhaustion, and strained working relationships, ultimately diminishing their overall wellbeing. NonLreporting was also widespread in our sample, with the most frequently cited reasons being fear of reprisals and the fact that the perpetrator was often a superior. It is essential that institutions implement measures to protect employees from sexual harassment as part of the broader stance against discrimination and violence in healthcare and protect those who report such incidents.

### Recommendations

Creating a safe and respectful healthcare environment requires urgent and sustained action. Healthcare institutions should implement comprehensive awareness programs, establish clear reporting pathways, and provide accessible support services for physicians and other healthcare workers. Policies should clearly state expectations regarding professional conduct and specify the consequences of violations. Also, training should be provided to help employees recognize sexual harassment and address common misconceptions, while supervisors and managers should receive training in conflict management, emotional regulation, and supportive communication to better assist individuals who may be reluctant to report harassment[46]. Evidence suggests that participation in sexual harassment prevention training can improve employees’ knowledge and ability to prevent and address harassment [47]. Furthermore, promoting gender equality in leadership positions, particularly in traditionally male-dominated specialties and departments, may help foster a more inclusive workplace culture and contribute to efforts to reduce sexual harassment[48, 49]. Lastly, Fear of reprisals should not continue to prevent victims of sexual harassment from reporting; therefore, zero tolerance for sexual harassment and accountability for perpetrators regardless of status are essential. Institutions must protect those who speak and safeguard them from retaliation, including job loss. International policy initiatives have emphasized the importance of preventing and addressing workplace sexual harassment. Key recommendations include mandatory anti-harassment training, clear reporting procedures, and ensuring that employees are informed about their rights and available support services[50]. Similar measures may help strengthen efforts to prevent and respond to sexual harassment within Swiss healthcare institutions.

## Data availability

The data cannot be publicly shared because the descriptive accounts of sexual harassment contain highly sensitive information that could potentially reveal participant identities even after deLidentification. Data may be made available to qualified researchers who meet the criteria for access to confidential data. Requests must be submitted for dual approval through the University of Lausanne Ethics Committee and the Institute of Humanities in Medicine with KTJD in copy. Also, as participants didn’t give their consent on the consent form for their data to be shared publicly, we cannot make it publicly available for this reason, as this is an obligation in Switzerland.

## Acknowledgements

We are grateful to the VSAO for helping us with the survey’s distribution and for their input during its creation. Additionally, we would like to express our gratitude to all the physicians who took the time to fill out the survey and to the staff at the Institute of Humanities in Medicine (UNIL - CHUV) who assisted with translation and verification of the questionnaire to guarantee that the survey was accessible in all Swiss languages.

## Authors contributions

All authors made significant contributions to the development of the article. KD conceptualised the idea of the study and the research methodology, took part in data analysis and interpretation, and wrote the first draft of the manuscript. RJJ participated in conceptualisation of the study, research methodology, editing the first draft of the manuscript and supervision of the research. PB participated in research methodology, edited the first draft of the manuscript and supervised of the research. All authors read and approved the final version of the manuscript.

## Supporting information

**S1 Table. Some descriptive free text responses on question: what action was taken by superior when you reported harassment**

