## Supplementary material for "Swiss Physicians’ Experiences Of Sexual Harassment At Work. A Nationwide Survey": S1 Table

**S1 Table. Some descriptive free text responses on question: what action was taken by superior when you reported harassment**

| Themes | Examples of Descriptive texts from respondents |
| --- | --- |
| Addressing the perpetrator | “Supervisor came into the emergency room and pointed out to the patient that sexual harassment would not be tolerated” |
| Discharging the patient | “Support with, for example, taking over patient communication or reprimanding the patient or discharging them” |
| Being told it was difficult to act | “She explained to me that it is difficult to take action against it. She even told a superior, but he dismissed it as “oversensitivity” |
| Being told to tolerate since they still needed help from their superiors | “They advised me to endure it in order to receive my documents signed by my boss. To understand that unfortunately I am not the first and will not be the last, and that it is difficult to fight alone against a ‘great professor’, stigmatisation, the lack of colleagues who will agree to testify, etc” |
| Being granted support by supervisors | “From then on, I was no longer assigned to the operating theatre where the person who had harassed me worked. That was what I wanted. I was given the option of reporting the incident to HR. However, I was told that I would have no chance of success and that it would damage my reputation. As I was still in training and was afraid of not being able to become a specialist, I changed jobs” |
| being encouraged by administrative staff to go public with case/direct confrontation | “The secretary told me to threaten to go public with my supervisor. After I confronted him, my supervisor stopped behaving as described, but then he just bullied me” |
| Documented in patients record | “this was documented in the patients medical record” |
| Mediation by superiors | “They organized a meeting individually with me and the person who harassed me” |
| Sending a male physician to see patient as way to prevent sexual harassment of females | Superior: There is no evidence to support this claim. Colleagues: It would be advisable to adjust behaviour, and if possible, send a male person to the patient |
| Refusal of care | patient was refused care |
| Exclusion of perpetrator from training | The colleague was excluded from the further training programme for doctors |
| Confrontation of perpetrators by senior colleagues | In the case of the junior assistants, the chief physician spoke to those involved. The junior assistants expressed their apology, at least in part, and subsequently displayed more appropriate behaviour, at least in part. A contact point for discrimination was established |
| Dismissal of perpetrator | “The individual in question was dismissed, as my report advocated for the reporting of future cases” |
| No action taken and perpetrator promoted instead | “The final result was that, despite the conflicting accounts, the individual in question was promoted to a higher position (from senior consultant to head physician), despite this not being the only instance of this nature”), |
| Sexual harassment being normalised | “I was advised to persevere in order to obtain the requisite documentation bearing the signature of my superior. It became apparent that I was not the first individual to encounter this situation, and I would most likely not be the last” |
